# Development of a Small, Low-Power Real-Time Phase-Dependent Neuromodulation System

**DOI:** 10.1101/2025.05.06.25327003

**Authors:** Kimberley Wyse-Sookoo, Toren Arginteanu, Nicolas Norena-Acosta, Matt Udvardy, Helena Ljulj, Kelly Mills, William Anderson, Yousef Salimpour

## Abstract

Neurological diseases and neuropsychiatric disorders are often characterized by abnormal neural oscillations, such as exaggerated synchronization or suppression within a narrow frequency band and complex oscillation coupling which disrupt normal brain function and contribute to debilitating symptoms. Phase-dependent stimulation (PDS) offers a promising solution by synchronizing electrical stimulation with specific phases of neural oscillations, thereby enhancing therapeutic precision and efficacy. However, the widespread clinical adoption of PDS is hindered by technological challenges, including the need for accurate detection and prediction of neural oscillatory phases in real-time, stimulation management, stimulus artifact removal, fast communication, and adaptable hardware for dynamic neural environments. This study aims to address some of these challenges by leveraging adaptive System-on-Chip and Field-Programmable Gate Array (FPGA) technology, which offers the computational power and flexibility required for real-time neural signal processing and management. Specifically, we propose to optimize, integrate, and validate our PDS technique within this advanced hardware framework to develop a unified, closed-loop phase-dependent neuromodulation system. We evaluated our device’s performance by assessing its latency and accuracy in targeting specific phases of stimulation on both simulated signals and intraoperative cortical and subcortical recordings. Our findings indicate that the device successfully sent stimulation commands in time with the occurrence of target phases with both high accuracy and low latency for extended time periods. This work has the potential to transform therapeutic approaches for disorders with well-described brain network dysfunction, offering a precise, adaptable, and safer alternative to traditional stimulation techniques.

## 1. Introduction

Abnormal neural oscillations are a characteristic of multiple neurological conditions. Among these conditions are movement disorders such as Parkinson’s (PD) [1-4] and Huntington’s disease (HD) [5], epilepsy [6], neuropsychiatric disorders such as schizophrenia (SZ) [7-13], and cognitive disorders such as Alzheimer’s disease [6, 14, 15]. These abnormalities may manifest as narrow-band synchronization or suppression. Individuals with PD often have excess beta band power in the subthalamic nucleus (STN) [3, 16]. Some studies have demonstrated that SZ patients have increased low frequency activity and suppressed high frequency activity at rest compared to healthy controls [7], and SZ patients who experience auditory verbal hallucinations (AVHs) have significantly increased beta band activity in the left inferior parietal region and left medial frontal gyrus compared to non-AVH SZ patients [17]. In HD, the most prominent electrophysiological abnormality is the suppression of alpha band activity [5]. Alzheimer’s and other cognitive disorders have been associated with abnormal levels of hippocampal [14, 15] and medial prefrontal [15], auditory, and visual cortex [14] gamma band power. Alternatively, there may be abnormal levels of cross frequency coupling. Elevated levels of beta-gamma phase amplitude coupling (PAC) have been observed in the motor cortex of individuals with PD [4], and increased theta-gamma PAC in the left temporal regions appears to correlate to the presence of auditory verbal hallucinations in SZ [11, 12].

Phase-dependent stimulation (PDS) is a stimulation paradigm that synchronizes electrical stimulation pulses with a specific phase of a neural oscillation. In doing so, PDS has the potential to augment or suppress the power of a select frequency band and/or modulate the level of synchrony between two frequency bands [18-20] at relatively low stimulation frequencies (<30 Hz). In PD, PDS has been able to modulate human motor cortex beta-gamma PAC [20] and STN beta band power [18], which both correlate to motor symptom severity [3, 4, 16], and has been shown to alter movement quality in animal models of PD [21, 22]. In a similar fashion, PDS has the potential to modulate theta-gamma PAC in the parahippocampal gyrus, which may alter performance in working memory tasks [19]. Many other neurological disorders that stem from abnormal neural oscillations [6, 14, 15, 19] may also be candidates for treatment by PDS.

However, the widespread clinical adoption of PDS is hindered by technological challenges. To our knowledge, there are no existing implantable devices that are able to consistently deliver stimulation in time with the occurrence of a specific oscillation phase. The main challenges in implementing a PDS device are minimizing the latency between phase detection and the delivery of a stimulus and avoiding the interference of stimulation artifacts in the recorded neural signals [18, 20, 23]. While autoregressive (AR) modeling can be used to effectively eliminate stimulus artifacts [20, 24, 25], existing PDS systems still struggle with minimizing computation latency enough to achieve consistent stimulation for more than a few hundred milliseconds [18, 21, 22].

In this study, we address this challenge by leveraging adaptive multiprocessor System-on-Chip (MPSoC) and Field-Programmable Gate Array (FPGA) technology, which offers the computational power and flexibility required for real-time neural signal processing and management. FPGA based devices are highly suitable for this task as they are capable of performing complex computations at very low latencies without consuming much power [26]. These devices have already been used for a number of real time applications in image processing [26, 27], deep brain stimulation (DBS) [28], diffuse correlation spectroscopy [29], and brain computer interfaces [30]. Additionally, FPGAs are easily reconfigurable and extremely compact [31, 32], making them ideal for translation from proof-of-concept applications to chronically implanted devices. The integration of a central processing unit (CPU) and FPGA onto a single chip to create an MPSoC further reduces communication latency between the control plane and the data plane [33] and opens up the potential for a device with parameters that can be easily tuned to optimize stimulation for users’ specific oscillation profiles.

We have optimized, integrated, and validated a novel strategy for delivering phase-dependent neuromodulation within an MPSoC framework to further the development of a unified, closed-loop phase-dependent neuromodulation system. Our system is capable of recording neural signals, estimating and predicting beta band phases, and triggering a stimulation pulse within microseconds, allowing for extended periods of accurate PDS. We tested our device’s accuracy and latency on a pure 20 Hz sine wave and on local field potential (LFP) recordings from the motor cortex and STN. We also measured the portion of the total target phases that were identified by the device.

## 2. Methods

### 2a. Participant and Ethics Statement

Neural data was recorded from two participants who gave written informed consent. Left motor cortex electrocorticography (ECoG) recordings were obtained from a male with PD. Right STN microelectrode recordings were obtained from a female with PD. Both participants were in their 70s at the time of surgery. The study protocol was approved by the Johns Hopkins Medicine Institutional Review Board (IRB00309062).

### 2b. Device Description and Experimental Setup

Figure 1 contains an overview of the device schematic and experiment setup. A more detailed block diagram of the MPSoC modules can be seen in Supplementary Figure S1. An oscilloscope and function generator were used for sinusoidal PDS experiments. Motor cortex experiments utilized a 128-channel data acquisition system (NSP – Neural Signal Processor) (NeuroPort, Blackrock Microsystems, Salt Lake City, UT) and a Blackrock CereStim macrostimulation device. A Neuro Omega system (Alpha Omega, Nazareth, IS) was used for data acquisition and reception of stimulation commands in the STN experiments. An operator laptop enabled a control interface with the software application running on the Ultra96-V2 MPSoC board (Avnet), which established a serial connection over an SSH connection. The rest of the components include a click mezzanine adapter (96Boards, Linaro Limited), and a 22 click ADC (Texas Instruments).

**Figure 1:**
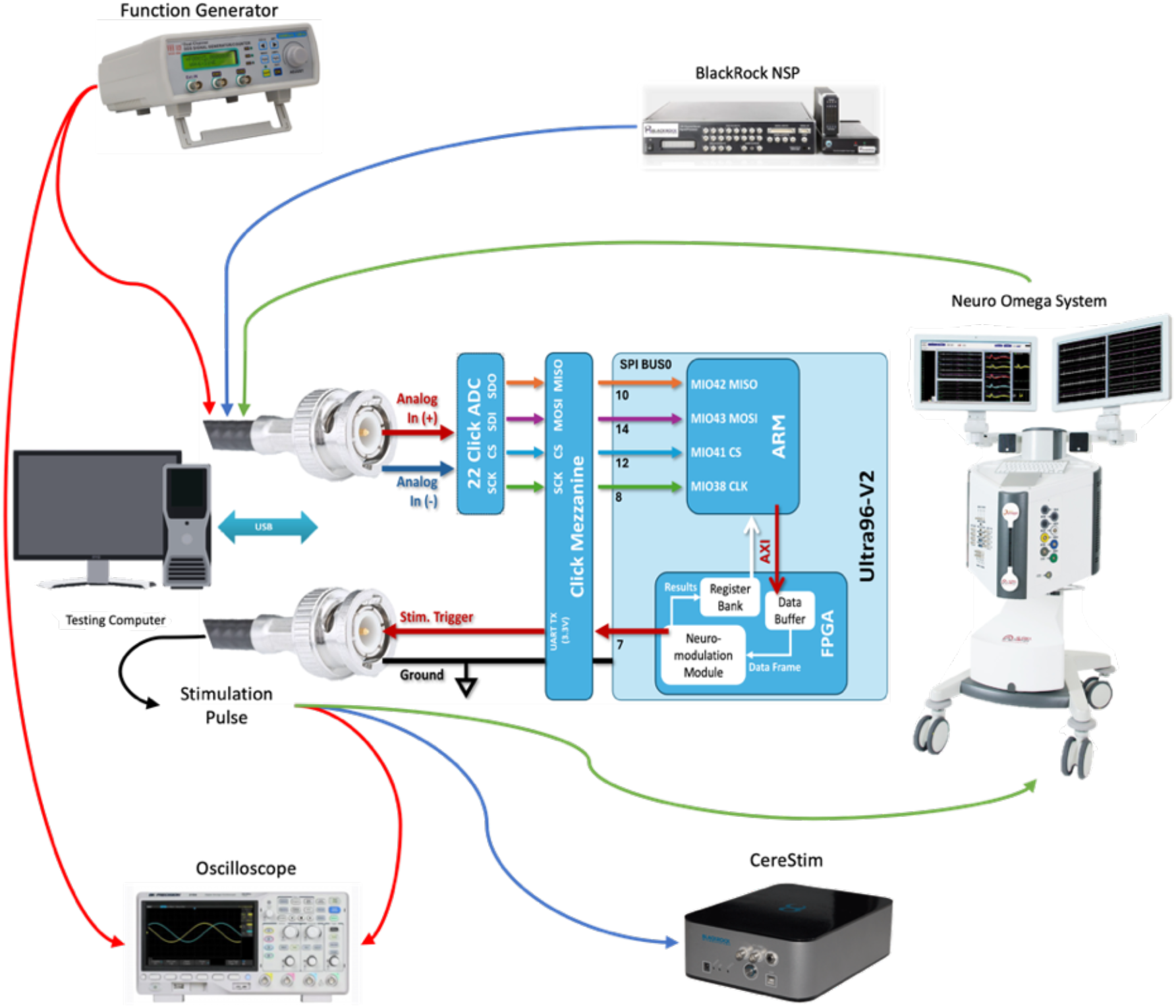
Block diagram of the device and hardware setup. Incoming neural data is passed through an analog to digital converter (ADC) and click mezzanine adapter board to be processed by the Ultra96-V2 multiprocessor system on a chip (MPSoC). The neuromodulation module of the programmed field programmable gate array (FPGA) hardware is responsible for bandpass filtering, extraction of instantaneous magnitude and phase, AR phase prediction, and sending stimulation commands. Red, blue, and green arrows represent hardware connections for the sine wave, motor cortex, and subthalamic nucleus (STN) experiments, respectively.

### 2c. Neural Recordings and Signal Preprocessing

Neural recordings were obtained while subjects were at rest during awake bilateral DBS implantation procedures. For motor cortex experiments, a 63-channel high density subdural ECoG grid (PMT Corp, Chanhassen, MN) was inserted through a burr hole and placed over the right primary motor and parietal cortices (Supplementary Figure S2). Motor cortex local field potentials (LFPs) were recorded by the Blackrock NSP at a sampling rate of 30 kHz. Amplification, down sampling to 1 kHz, and initial band pass filtering (1 – 250 Hz) were all performed by the NSP. STN LFP recordings were obtained by a Neuro Omega microelectrode and data acquisition system at a sampling rate of 44 kHz. The Neuro Omega system performed initial amplification, down sampling to 1375 Hz, and 300 Hz low pass filtering. Analog signals were reconstructed from the downsampled data to be sent to the MPSoC device.

### 2d. Neuromodulation Module

After preprocessing, data was sent through an analog to digital converter (ADC, 1 kHz sampling rate) and click mezzanine adapter board to the MPSoC neuromodulation module detailed in Figure 2. This module allows the device to filter the data, obtain instantaneous phase information, make AR guided predictions of when the next targeted phase will occur, and time simulation pulses accordingly. Beta band power was extracted by a 13-30 Hz finite impulse response (FIR) band pass filter in both the forward and reverse directions with zero phase distortion. Next, a Hilbert transform was applied to the data. To minimize edge effects, prior to the Hilbert transform, forecasted future data estimated using an AR model was appended to the data; the best-fit AR coefficients were estimated using the data recorded during a baseline period before stimulus began. The Hilbert transform returns a complex number from which the instantaneous frequency and phase of the signal were obtained and used to predict the timing of upcoming target phases (phase projection). These predictions are passed to the stimulation control block to plan future stimulation pulses. In parallel to phase projection, the frequency domain magnitude information from the Hilbert transformed data was sent to a thresholding block. The thresholding block disables stimulation if the beta band power drops below the set value in order to avoid stimulating based on non-beta band signals. A hold parameter equal to the length of one stimulation pulse was used to prevent subsequent stimulation pulses from being delivered before a prior pulse was completed.

**Figure 2:**
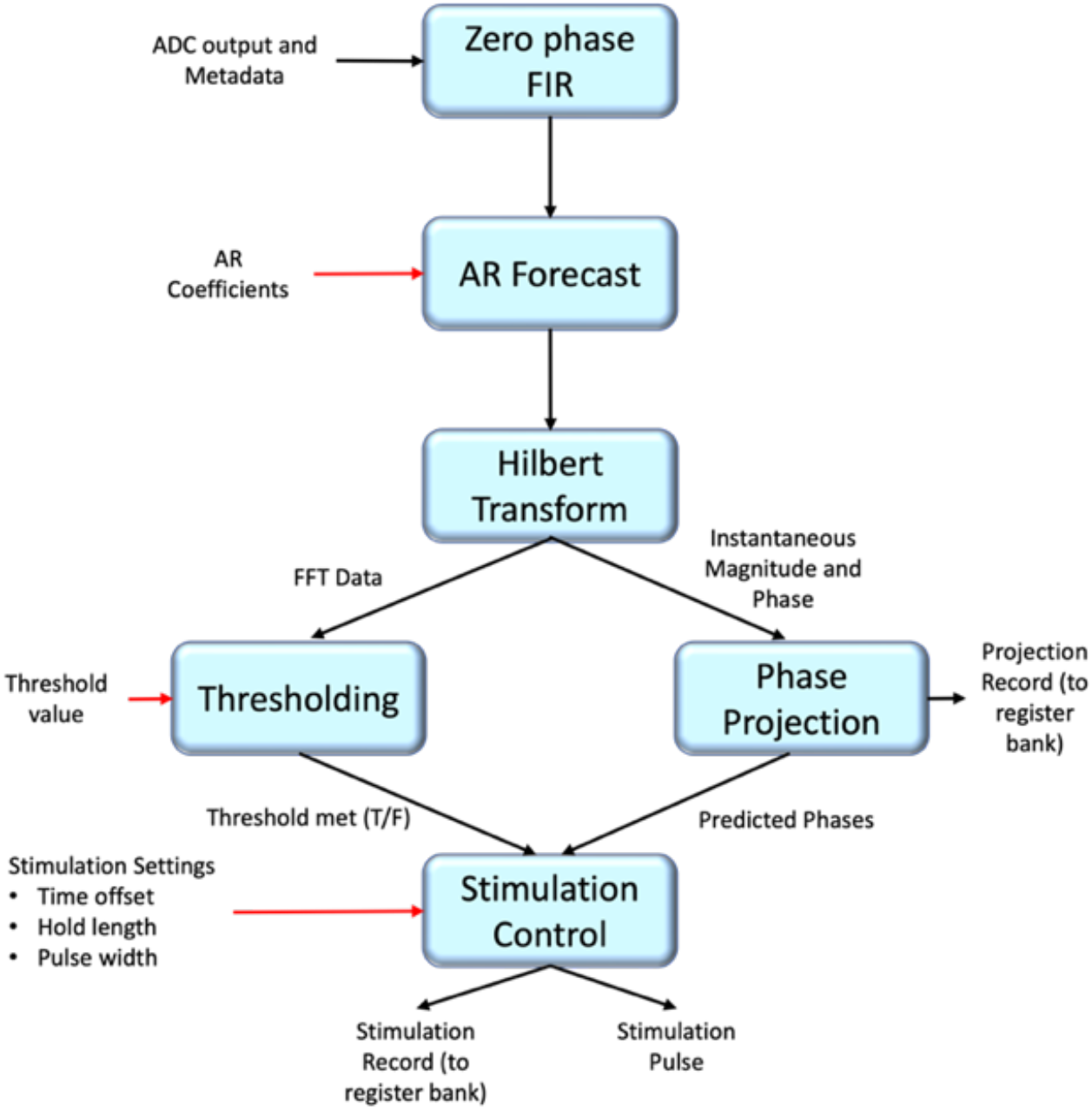
Block diagram of the neuromodulation module. Data enters the module from the ADC and is band pass filtered by the zero-phase finite impulse response (FIR) filtering block. The autoregressive (AR) forecast block appends forecasted future data to the filtered data to minimize edge effects in the Hilbert Transform block. The fast Fourier transform (FFT)-based Hilbert transform block produces FFT magnitude data, used for thresholding, and complex time-domain samples with easily computable phase which are used to determine the timing of stimulation pulses. The thresholding and phase projection blocks work in parallel to inform stimulation control. The thresholding block determines whether beta band power is dominant in the input signal, as a safeguard against stimulating based on noise or other frequency bands. The phase projection block predicts the time of upcoming target phases and passes these predictions to the stimulation control block which sends out stimulation commands. User defined inputs are denoted by red arrows.

### 2e. Sinusoidal phase dependent stimulation

The device’s accuracy was first tested on a pure 20 Hz sine wave with 500 mV peak-to-peak amplitude provided by a function generator. Peak phases, defined as the points where the instantaneous phase reached 0 degrees, were targeted with recorded stimulation pulse commands running for 360 s. The phase of the signal at the time of each stimulation pulse command sent from the device was calculated from the Hilbert transform of the input signal to evaluate stimulation accuracy. Stimulation consistency was estimated by the percentage of targeted peaks, which was calculated by dividing the number of stimulation pulse commands by the true number of peak phases in the input. Interevent intervals for the input signal peaks and the stimulation pulse commands were also calculated to verify that the stimulation frequency aligned with the frequency of the input signal. This process was then repeated on a sine wave with a frequency sweeping between 15 – 25 Hz every 5 s.

### 2f. Motor Cortex beta band phase dependent stimulation

Neural signals were recorded and processed as described above during awake bilateral DBS implantation procedures while the subject was at rest. Peak targeting stimulation timing was performed on incoming data from one channel that was determined by analysis of ECoG physiology to be over the motor cortex in real time for 500 s. To avoid interference from stimulation artifacts, stimulation commands sent from the MPSoC device to the CereStim were recorded, but no stimulation was delivered to the participant. Stimulation accuracy and consistency and interevent intervals were calculated as described in section 2e for the 13-30 Hz bandpass filtered input signal. In the rare occurrence that two pulses were triggered within less than 30 mS of each other, they were counted as one pulse, as our stimulation hardware is set to ignore subsequent pulses within this window to keep the stimulation frequency low. The average packet processing time was also recorded for the neuromodulation module on a portion of the recorded data.

### 2g. STN beta band phase dependent stimulation

To demonstrate that our device can be used in multiple brain regions, 35s of peak targeting stimulation were performed on LFP data recorded from a microelectrode in the STN. The data was then analyzed in the same way as in the motor cortex experiments (section 2f).

## 3. Results

### 3a. Sinusoidal PDS

Peak targeting stimulation was first delivered to a 20 Hz sine wave for a total of 360 s. The device was able to target 99.99% of all peak phases in the input signal, with 99.99% of the stimulation triggers occurring within 60º of a peak phase (Figure 3a). Next, the input was changed to a sine wave with a frequency varying linearly over time from 15 – 25 Hz to more closely mimic the natural shifts in the dominant beta band frequency that occur with different movement states [34]. PDS was delivered to this input signal for 135 s. When the stimulation threshold was met, 97.6% of the input peak phases triggered a stimulation command, with 90.9% of the stimulation commands occurring within 60º of a peak phase (Figure 3b).

**Figure 3:**
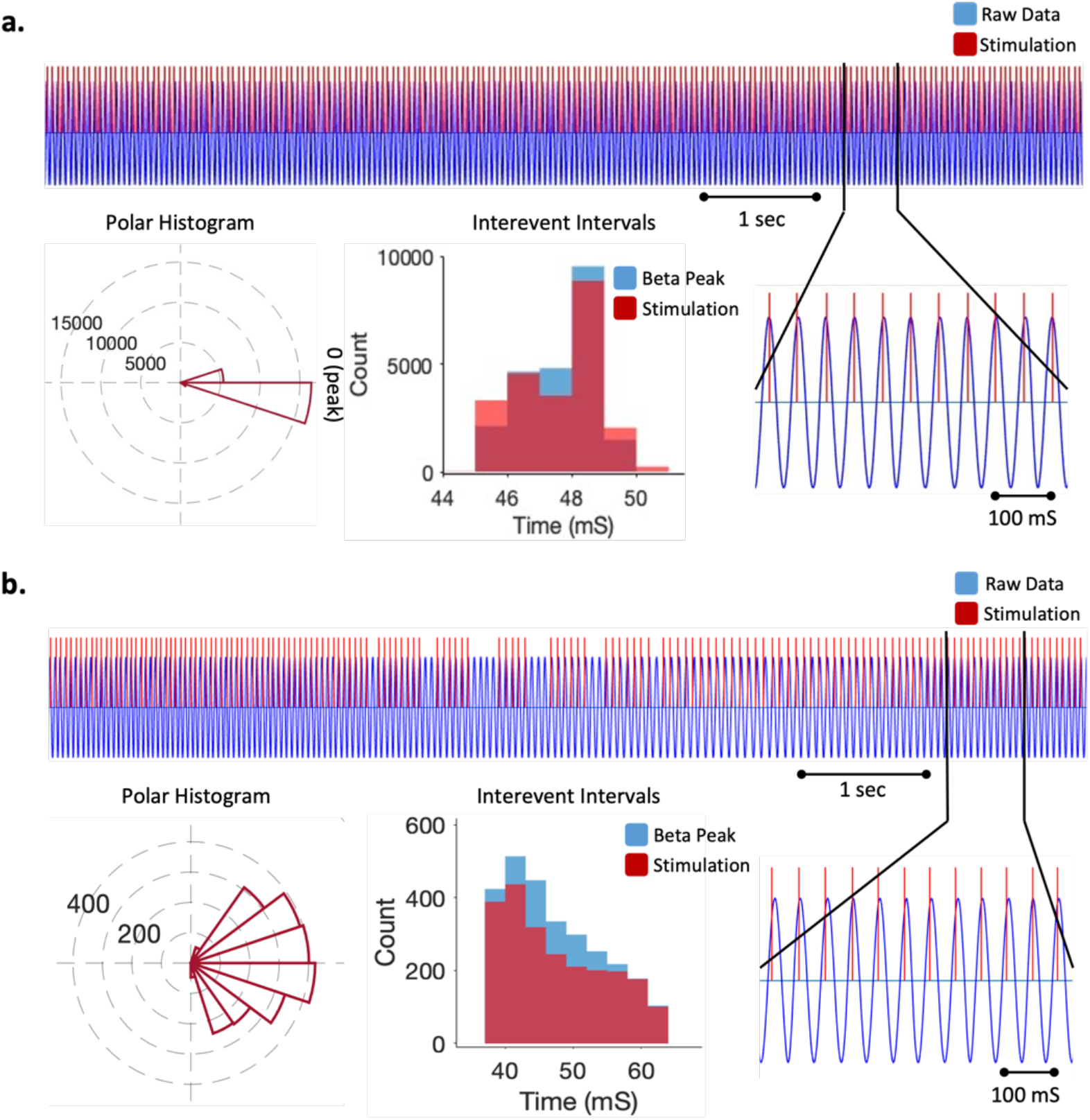
Results of sine wave stimulation testing. For each input type (**a**. 20 Hz stimulation; **b**. 15-25 Hz frequency sweep), a portion of the time series data is shown with the input signal in blue and stimulation triggers denoted by red vertical lines. Polar histograms depict the phases of the input signal during all stimulation triggers. The similarity of the interevent intervals for the peaks occurring in the input signal and the time between stimulation pulses is shown by overlapping histograms.

### 3b. Motor Cortex PDS

Peak targeting stimulation was delivered to incoming motor cortex data for a total of 500 s. The stimulation threshold was met for multiple epochs lasting between 2 and 13 seconds. While the stimulation threshold was met, 95.3% of the signal peaks triggered a stimulation command, and 68.0% of the stimulations were within 60 degrees of a peak phase (Figure 4). For a portion of the data, packet processing times for the neuromodulation module were able to be recorded. The average latency between phase projection results was 96.4 µs.

**Figure 4:**
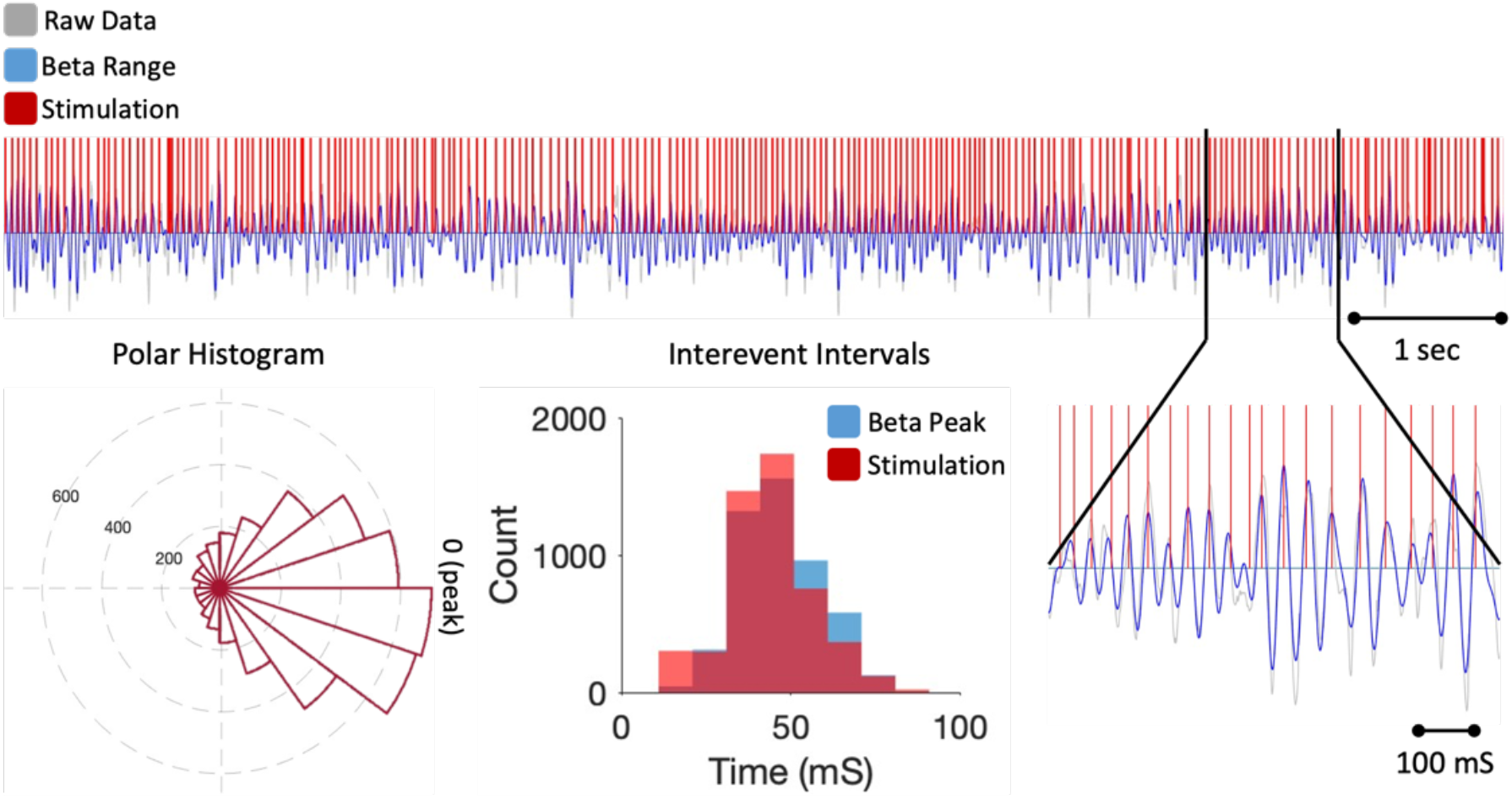
Results of motor cortex phase dependent stimulation (PDS). A portion of the data recorded during motor cortex beta-band peak targeting PDS is shown with the input signal in gray, the beta-band filtered signal in blue, and stimulation triggers denoted by red vertical lines. Polar histograms depict the phases of the input signal during all stimulation triggers. The similarity of the interevent intervals for the peaks occurring in the input signal and the time between stimulation pulses is shown by overlapping histograms.

### 3c. STN PDS

Peak targeting stimulation was delivered to incoming STN data for a total of 35 s. The stimulation threshold was met for the entire recording. 89.9% of the signal peaks triggered a stimulation command, and 60.0% of the stimulations were within 60 degrees of a peak phase (Figure 5).

**Figure 5:**
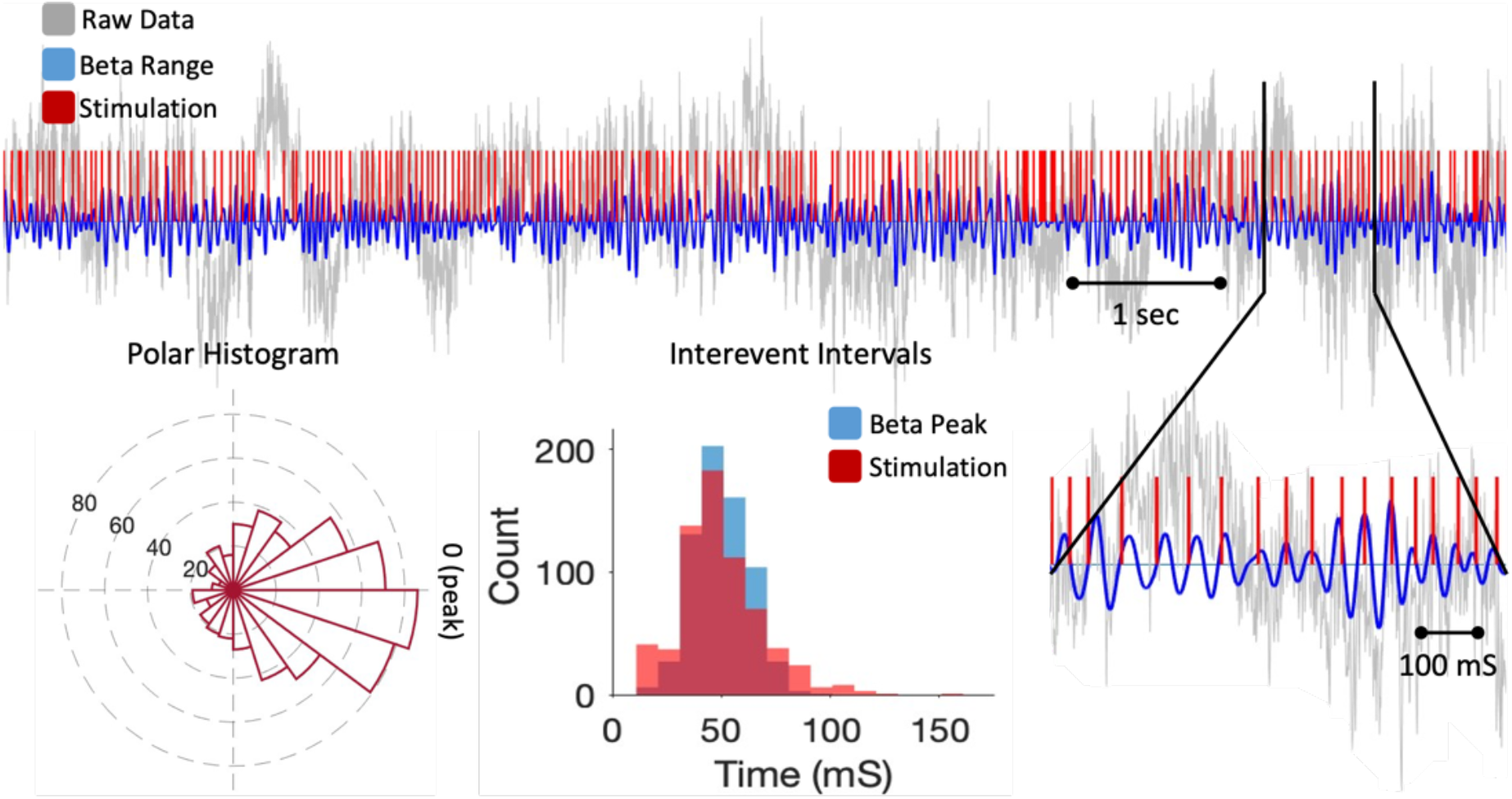
Results of subthalamic nucleus (STN) phase dependent stimulation (PDS). A portion of data recorded during STN beta-band peak targeting PDS is shown with the input signal in gray, the beta-band filtered signal in blue, and stimulation triggers denoted by red vertical lines. Polar histograms depict the phases of the input signal during all stimulation triggers. The similarity of the interevent intervals for the peaks occurring in the input signal and the time between stimulation pulses is shown by overlapping histograms.

## 4. Discussion

This work presents a novel method for using a low SWAP platform to deliver consistent phase-dependent stimulation during delimited analysis windows. Our proposed system’s increased stimulation timing stability and decreased computational latency compared to prior approaches make it a significant step towards a fully implantable system for the delivery of PDS. Minimizing device latency is a key factor in achieving consistent, on target stimulation. By taking advantage of the extremely low computational latencies of an FPGA, our device was able to predict and target upcoming peak phases with sub-millisecond processing times. Previous systems have only been able to achieve consistent PTS for a few hundred milliseconds at a time [18, 21, 22]. Here, we have shown that our device is capable of triggering consistent, on or near-target stimulation in real time for up to 360 s.

The majority of the stimulation triggers occurred near the targeted phase for all applications and our motor cortex phase targeting accuracy is very similar to that of previous studies applying PDS to the motor cortex with a PC-based system [20, 35, 36]. However, stimulation accuracy does appear to be affected when the input signal deviates from a pure sinusoid. We believe that this is due to the current system having static AR coefficients that are set before stimulation begins. If the center frequency of the beta band signal shifts from the center frequency of the baseline data used to set the AR coefficients, this could lead to less accurate predictions. In future iterations of the device, we will aim to dynamically alter the AR coefficients based on the evolving signal characteristics.

The effects of PDS on beta band power become more pronounced with a larger number of consecutive on or near target stimulations [18]. Overall targeting consistency remained quite high across sinusoidal, motor cortex, and STN testing, with a very low rate of missed target phases. This is supported by the similarity between the interevent interval histograms for input peaks and stimulation pulses (Figure 3-5). Although they do not state the percentage of phases that were correctly targeted by their systems, we believe our system’s consistency is comparable to that depicted during the high performing stimulation epochs in prior studies [20, 21]. A very small percentage of the stimulations that were triggered occurred at a frequency above 30 Hz. This would not pose a significant risk to the patient while actively delivering stimulation, as our stimulation hardware is programmed to stay below a predefined stimulation frequency, so these higher frequency pulses would not be delivered to the patient.

Although our motor cortex testing saw the device being used for the longest consecutive period of time, the stimulation threshold was only met for smaller epochs within the session. Our current system only allows for the threshold to be set at the beginning of a testing session. Further exploration is needed to determine an optimal threshold value, which may very likely be patient specific. Too low of a threshold may begin to trigger false positives, leading to excess stimulation triggers, while too high of a threshold could make stimulation too sporadic. Additionally, beta band spectral characteristics are known to change based on movement states [34], so it would be beneficial to incorporate functionality to more frequently update the threshold value based on spectral information rather than depending on a user-defined value.

While our results are promising, we recognize that this study is not without its limitations. Experiments were limited by the time and resource constraints of the operating room, and we were not able to test behavioral or PAC responses to stimulation since the device does not yet have artifact removal fully implemented. Future studies will aim to incorporate real-time artifact removal, evaluate this device on a more chronic timescale and over a larger set of subjects, and include analyses of repetitive movement tasks. While implementing artifact removal and making AR coefficients and thresholds dynamic may improve stimulation accuracy and consistency, further studies are necessary to determine whether these changes could be implemented without worsening device latency or necessitating online dynamic model coefficient adjustment, which could negate the initial benefits in latency seen with the current device.

## 5. Conclusion

Our device directly addresses the issue of minimizing latency within PTS systems, allowing for long periods of consistent stimulation in both cortical and deep brain structures. Future studies will focus on incorporating stimulus artifact removal capabilities and dynamic AR coefficients and evaluating device performance while stimulation pulses are being delivered to the cortex. We believe that the development of this device is an important step towards creating a chronically implantable PTS system.

## Data Availability

All data produced in the present study are available upon reasonable request to the authors.

## Acknowledgement

The authors express sincere gratitude to our participants, surgical team, and the JHU Neuromodulation and Advanced Treatments Center.

