## Supplementary Figures for "Development of a Small, Low-Power Real-Time Phase-Dependent Neuromodulation System"

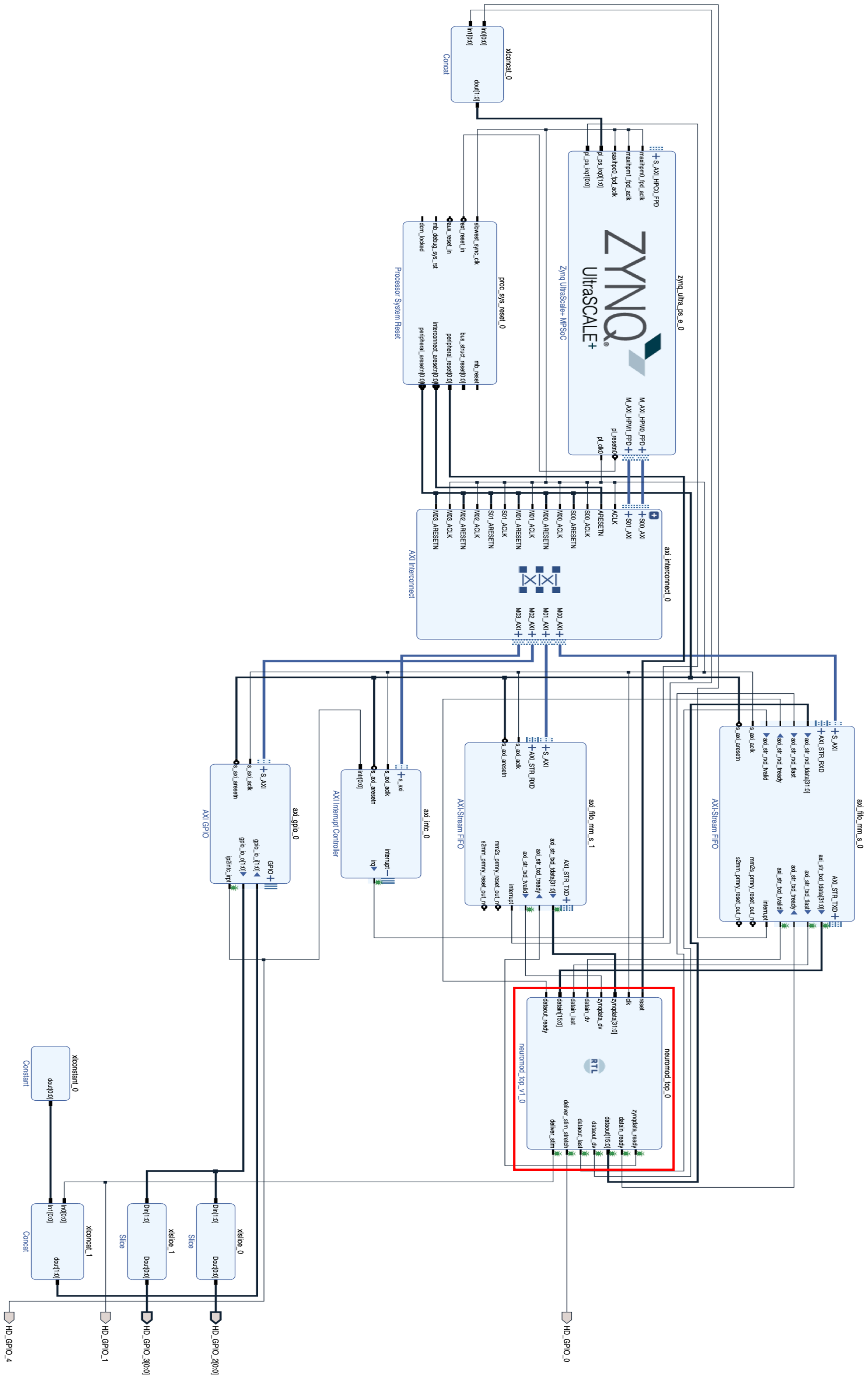


Figure S1: Detailed block diagram of the MPSoC modules. The neuromodulation module is outlined in red.

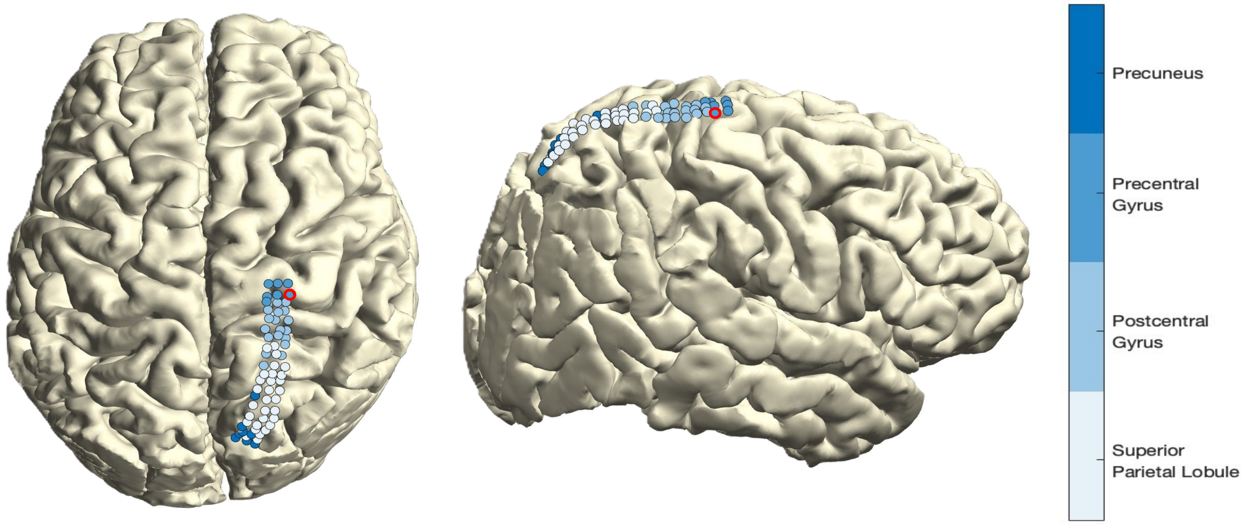


Figure S2: Reconstruction of preoperative MRI with electrode locations. MRI reconstruction was performed in Freesurfer (version 7.4.1, http://surfer.nmr.mgh.harvard.edu/) and co-registered with intraoperative CT data using the FieldTrip toolbox for MATLAB ([37]; version 20231113, http://fieldtriptoolbox.org, Donders Institute for Brain, Cognition and Behaviour, The Netherlands). Anatomical locations of the electrodes were determined using a combination of the Brainetomme and MNI atlases. The recording contact is outlined in red.
